# Changing dynamics of SARS-CoV2 B.1.617.2 (Delta variant) outbreak in the United Kingdom: Shifting of SARS-CoV2 infections from younger to elderly populations with increasing hospitalizations and mortality among elderly

**DOI:** 10.1101/2022.03.05.22271084

**Authors:** Venkata R. Emani, Abirath S. Nakka, Kartik K. Goswami, Shaila R. Emani, Kailash R. Maddula, Nikhila K. Reddy, Nidhi K. Reddy, Dheeraj Nandanoor, Sanjeev Goswami, Raghunath Reddy

## Abstract

**Background:** To assess the comprehensive dynamics of outcomes during the SARS-CoV2 B.1.617.2 (Delta variant) compared to the Alpha variant outbreak in the United Kingdom.

**Methods:** In this observational study of the cases reported by Public Health England for confirmed (sequencing and genotyping), SARS-CoV2 cases Delta variant (n=592,692) and Alpha variant (n=150,934) were used. Outcomes were analyzed by age groups and compared with all reported weekly cases in the UK.

**Results:** The Delta variant surge is associated with a significantly lower case fatality rate (0.43% vs 1.07; RR 0.39; 95% CI 0.37-0.42; P<0.0001); lower odds of hospitalization (2.1% vs 3.0%; RR 0.70; 95% CI 0.68-0.73; P<0.0001) than the Alpha variant. During the Delta variant surge there were significant increased cases (11.3% to 21.1%, RR 1.87; 95% CI 1.84-1.89; P<0.0001), hospitalizations (40.2% to 56.5%; RR 1.40, 95% CI 1.3-1.46; P<0.0001) among confirmed Delta variant cases in the ≥50 years age group during the August 3-September 12, 2021 period compared to earlier reported cases. There was also a significant increase in total weekly COVID-19 deaths noted among ≥70 years old age group (71.4% to 75.1%; RR 1.05; 95% CI 1.01-1.08; P=0.0028) during August 6-October 8, 2021 compared to June 4-July 30, 2021 period.

**Conclusions:** The Delta variant surge is associated with significantly lower mortality and hospitalizations than the Alpha variant. As the Delta variant surge progressed, ≥50 years old had a significant increased percentage of cases, hospitalizations and a significant increased COVID-19 deaths occurred among ≥70 years old age group.

## INTRODUCTION

The Coronavirus disease 2019 (COVID-19) global pandemic caused by SARS-CoV-2 (severe acute respiratory syndrome coronavirus 2) began in early 2020 and the mutant variants of SARS-CoV2 are continuing to drive the pandemic.

The SARS-CoV2 B.1.1.7 (Alpha variant) was first detected in a sequence from UK in December 2020 (VOC-20DEC-01) and it became the predominant variant causing widespread transmission in the UK until late April 2021^1,2^. The SARS-CoV2 B.1.617.2 (Delta variant) was reported initially in India around in December 2020, and was later identified in the UK as a variant of concern on May 6, 2021 (VOC21APR-02)^3,4^. Until then about 196,341 cases of Alpha variant were reported with 4009 deaths in UK as of UK technical briefing 10 dated May 7, 2021^4^. Since then, the proportion of B.1.617.2 Delta variant cases sequenced on a steadily increased and the secondary attack rates were found to be higher for VOC-21APR-02 (B.1.617.2) than for VOC-20DEC-01 (B.1.1.7) in travelers and non-travelers as of UK technical briefing 12, dated May 22, 2021^5^. The Public Health England also reported that 74% of sequenced cases and 96% of sequenced and genotyped cases are Delta and the early data from both England and Scotland demonstrated an increased risk of hospitalization with the Delta compared to the Alpha variant by briefing 15 on June 11, 2021^6^. By briefing 16 as reported on June 18, 2021, the Delta variant comprised 91% of sequenced cases and among cases that completed the 28-day follow-up period, the crude case fatality rate was noted to be lower for Delta than other variants^7^.

As of September 12, 2021, the Public Health England reported 227, 214 cases of the Alpha variant with 4,353 deaths and a total of 593,861 Delta variant cases with 2,547 deaths^8^. There were reports of increased cases and hospitalizations of the younger population with the Delta variant compared to prior variants including the Alpha variants^6,9–13^. However, there were troubling signs of increased case fatality rate with increased hospitalizations noted among reported cases of the Delta variant especially in over 50 years old age group though UK technical briefings 17-23^8,14–19^. The outcomes of the Delta variant on comparison with Alpha variant and the outcomes of Delta variant among age groups is not completely understood.

We performed a comprehensive analysis of the confirmed (sequencing and genotyping) Delta variant cases, hospitalizations, and deaths among age groups <50years and ≥50 years and compared them with outcomes of the confirmed (sequencing and genotyping) Alpha variant. We also performed a comprehensive analysis of trends on percentage of all cases, hospitalizations, and deaths among various age groups during the Delta variant surge and compared them with prior surges particularly with the Alpha variant to understand the reasons for causes of hospitalizations and mortality.

## METHODS

The SARS-CoV2 B.1.617.2 (Delta variant) initially reported in India around December 2020, has since became the dominant strain during the recent outbreaks in various countries including UK^3,6^.

In this observational study, we analyzed the nationwide data of the confirmed (sequencing and genotyping) Alpha and Delta variants SARS-CoV2 cases as reported by the Public Health England between February 1 to September 12, 2021. We also analyzed for any changes in all reported weekly SARS-CoV2 case rates, hospitalizations, and deaths among various age groups during the Delta variant surge in UK and compared them with prior surges (July 5, 2020 to October 8, 2021).

### Analysis of the Confirmed Delta variant and Alpha variant outcomes

We analyzed the data of 592,692 confirmed Delta variant cases and 150,934 confirmed Alpha variant cases reported by Public Health England in between February 1-September 12, 2021 through their technical briefings^8,14–19^. We performed an analysis of overall and sub-group outcomes; odds of hospitalization (hospitalizations/cases), hospitalization-death rate (deaths/hospitalizations), and case fatality rates (deaths/cases) among age groups <50 years and ≥50 years of all confirmed Delta variant cases for each reporting period during June 21-September 12, 2021. The comparison of outcomes in between the Delta variant and Alpha variant was performed among all confirmed cases reported for the period ending September 12, 2021. The Public Health England stopped reporting the Delta variant outcomes among age groups, <50 years and ≥50 years after briefing 23 (for period ending September 12, 2021; the week 36) and we are unable to further analyze the data on confirmed cases of Delta variant since September 12, 2021 reporting period.

### Study of demographic shifts during the Delta variant surge and correlation with all weekly reported SARS-CoV2 outcomes in UK

An initial analysis was performed to assess the distribution of confirmed Delta variant cases, hospitalizations and deaths among age groups <50 years and ≥50 years in each reporting period starting from period ending June 21, 2021 (briefing 18) to September 12, 2021 (briefing 23) to evaluate for any changes in the outcomes among the studied age groups. The changes in outcomes (odds of hospitalization, hospitalization-death rate and case fatality rates) of the confirmed Delta variant cases for each reporting period (June 21-September 12, 201) among all ages, <50 years and ≥50 years old was determined.

We performed further analysis of all confirmed Delta variant cases, hospitalizations, and deaths reported as of June 21, 2021(briefing 18) and all the new confirmed Delta variant cases, hospitalizations, and death outcomes reported in between each briefing from June 21, 2021(briefing 18) through September 12, 2021 (briefing 23). We analyzed to determine if there any temporal changes in the percentage of new confirmed cases, hospitalizations and deaths among <50 years and ≥50 years age groups during the course of the Delta variant surge in between each reporting period on comparison with the immediate prior reporting period (from June 21-September 12, 2021).

We also analyzed the available data on all nationwide weekly reported SACRS-CoV2 cases, hospitalizations and deaths in UK. We used Public Health England’s database (National flu and COVID-19 surveillance reports) to analyze the weekly changes in proportion of all reported SARS-CoV2 case rates and hospitalization rates per 100,000 population among various age groups since July 5, 2020 (week 27) through October 8, 2021 (week 40)^20^. The UK Office of National Statistics dataset was used to analyze the percentage of all reported COVID-19 weekly deaths among same age groups of SARS-CoV2 case rates in National flu and COVID-19 surveillance reports for same period (week 27, 2020 to week 40, 2021)^21^. The demographic shift in outcomes among the confirmed Delta variant cases (February 1-August 2, 2021 and August 3-September 12, 2021) and the total weekly deaths among demographic age groups during the Delta variant surge were also compared (Table 4). The weekly reported trends of case rates and hospitalizations per 100,000 specified age group population; case rates per 100,000 specified age group population and percentage of total COVID-19 deaths among various age groups were compared from July 5, 2020 to October 10, 2021 by plotting of the analyzed data on Figures 1-4.

**Figure 1:**
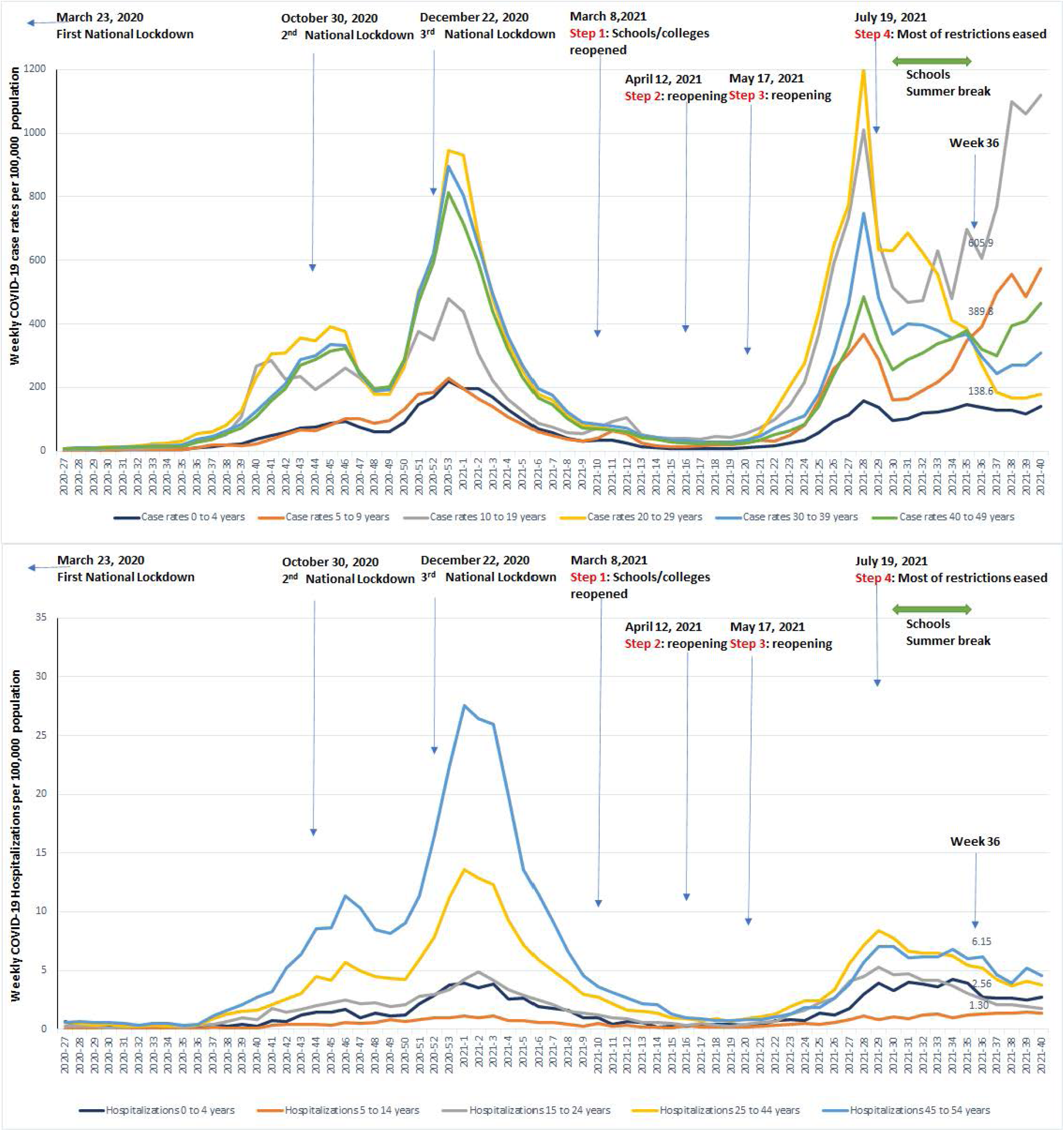
The upper graph shows SARS-CoV2 weekly case rates per 100K population. The case rates for 0-4 years, 30-39 years and 40-49 years are lower during the Delta variant surge than prior surge. The case rates for 5-9 years, 10-19 years are 2-3 times higher, and a slight increase in case rates among 20-29 years during the Delta variant surge. Significantly increased cases (more than prior surges) among 5-9 years and 10-19-years age groups were associated with Step 1 of March 8, 2021 school/colleges reopening, with decreased cases among same ages noted during school summer break. Lower graph shows hospitalization rate per 100K population among age groups listed, with hospitalizations peaking during Delta variant surge (29^th^ week in 2021) and gradual decline since then during the Delta variant surge until studied period ending 40^th^ week in 2021. Despite very high cases rates/100K among 5-9 years, 10-19 years and 20-29 years age groups during the Delta variant surge, the hospitalization rates among 5-14 years, 15-24 years age group during Delta variant surge are the same as the last surge and the 25-44 years and 45-54 years age groups showed lower hospitalization rates during Delta variant surge. This data is correlating well with table 4 data on under 50 age group for the period ending week 2021-36, showing decreased percentage of cases and hospitalizations over time during the Delta variant surge. The overall lower rates of hospitalizations among all age groups during the Delta variant surge than the prior surge is correlating well with the data on table 3 for odds of hospitalization.

**Figure 2.**
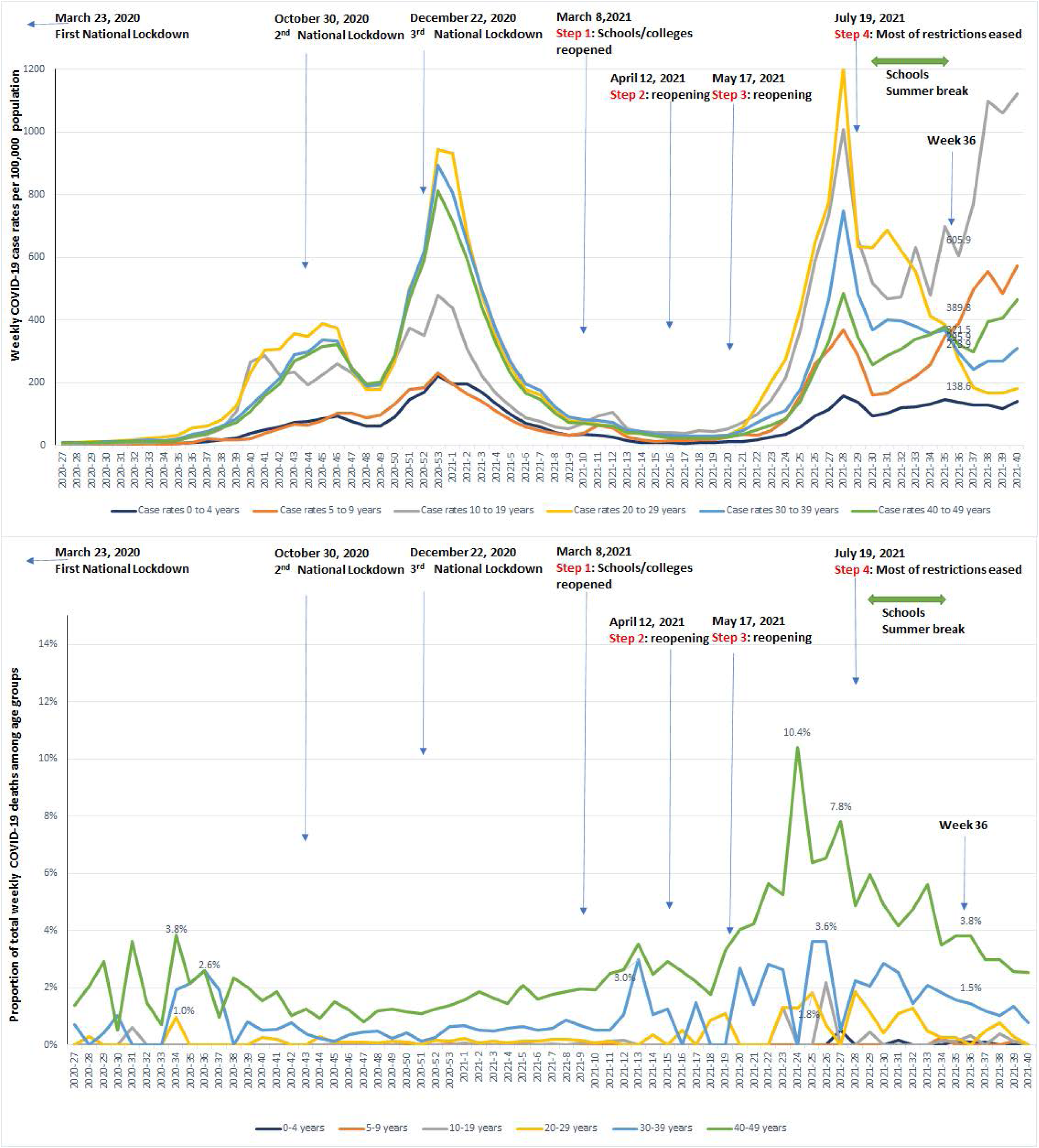
The upper graph shows SARS-CoV2 weekly case rates per 100K population, lower graph shows the percentage of weekly COVID-19 deaths among similar age groups. Despite higher proportion of cases among 5-9 and 10-19-years old age groups during the Delta variant surge, very low percentage of total deaths noted during Delta variant surge among 5-19 years relative to number of increased cases. Increased deaths among 20-29 years age correlated with similar increase in cases during the Delta variant surge. There was an increase in the percentage of deaths during Delta variant surge (weeks 22-33 in 2021) among 40-49 years old (despite decreased cases during Delta variant surge than before) and came back to same baseline of <4% noted during 27-25 weeks in 2020. *All age groups under 50 years showed downward trend in mortality during the Delta variant surge as reflected by the percentage of weekly deaths after peaking around 24-27 weeks in 2021 during the beginning of the Delta variant surge*.

**Figure 3:**
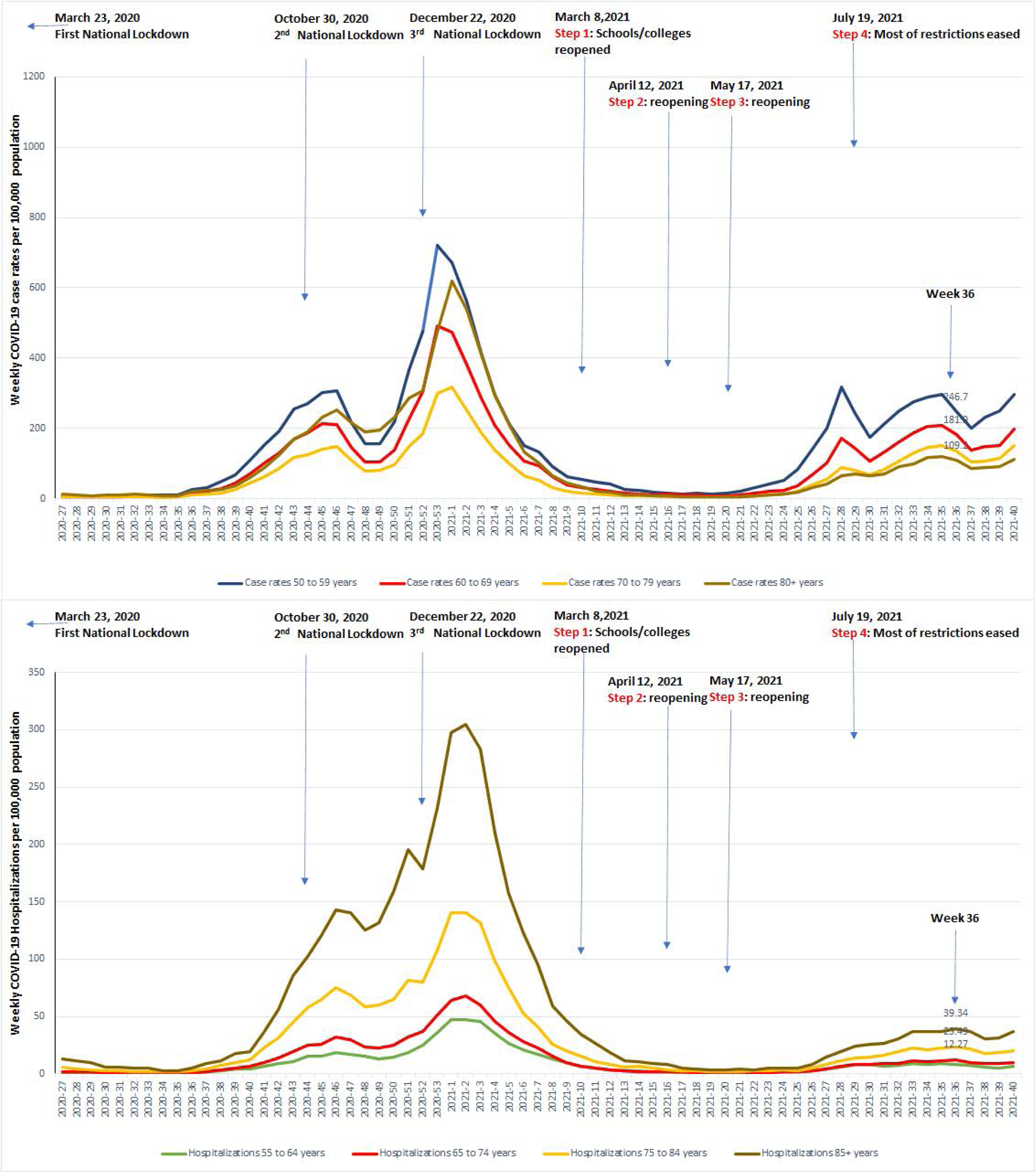
The upper graph shows SARS-CoV2 weekly case rates per 100K population. The cases rates/100K for over 50 years age groups much lower during the Delta variant surge than the prior surge. However, there is steadily increased cases among all age groups over 50 years old during the Delta variant surge, without any decline that was seen in under 50 years age groups in figures 1-2. The lower graph shows hospitalization rate per 100K population among age groups listed, with hospitalizations also steadily raising among over 50 years age groups (hospitalizations particularly among over 70 years age group are much lower during Delta variant surge than prior surge), this trend is similar to the trend that was seen on the Table 4 data regarding increased percentage of cases hospitalized among over 50 years age group. The overall lower rates of hospitalizations among all age groups during the Delta variant surge than the prior surge is correlating well with the data on table 3 for odds of hospitalizations.

**Figure 4:**
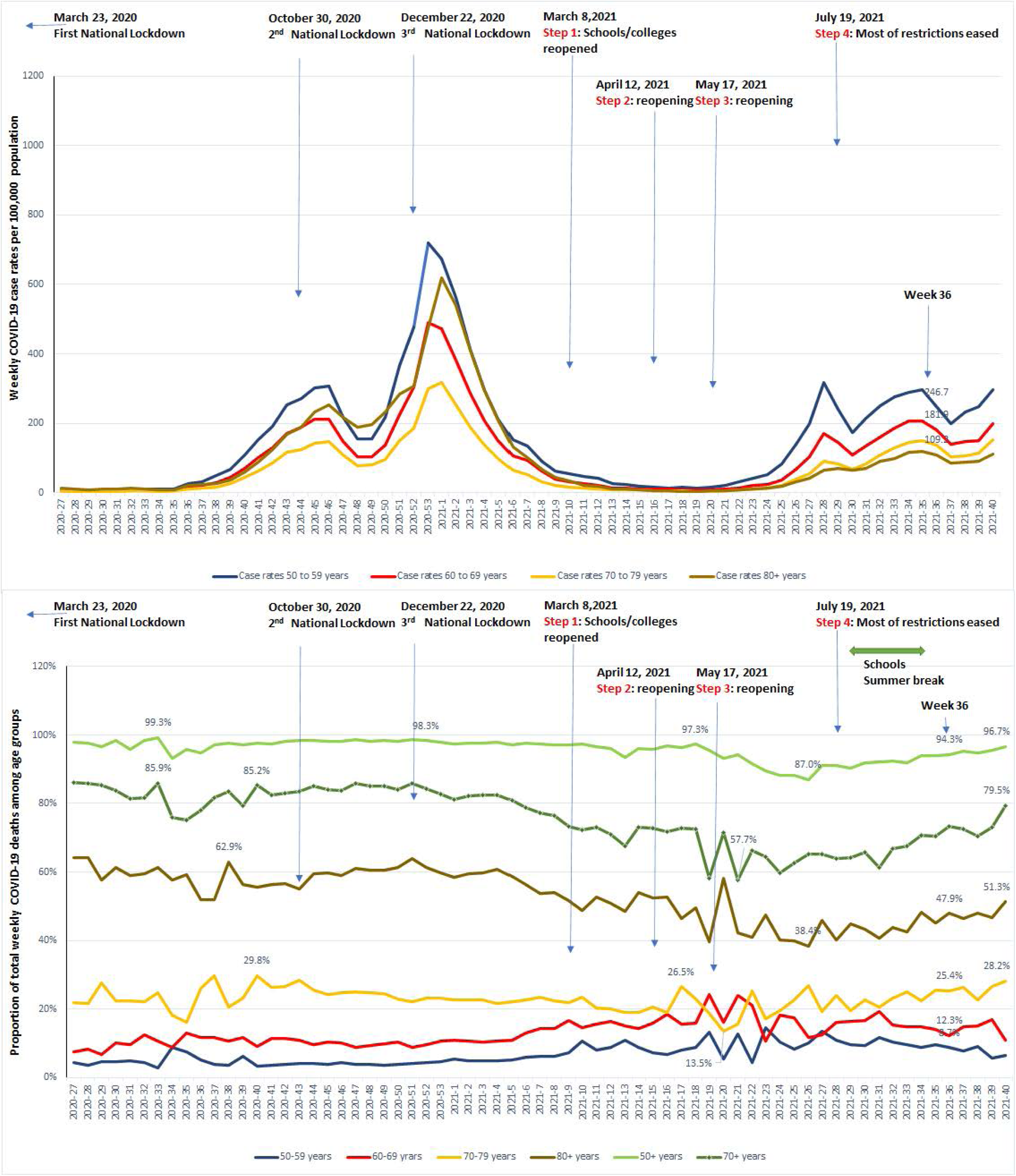
The upper graph shows SARS-CoV2 weekly case rates per 100K population, lower graph shows percentage of weekly COVID-19 deaths among similar age groups over 50 years. All age groups over 50 years showed decreased percentage of deaths during initial phase of Delta variant surge (weeks 24-30 in 2021), an increased percentage of deaths among 70+ years since then until study period ending week 40 in 2021. The percentage of deaths among over 50 years showed decline during weeks 22-27 during Delta variant surge, since then the percentage of weekly deaths came back to >95% same as prior to Delta variant surge with most of this increase is due to deaths in population over 70 years old.

### Statistical analysis

The comparison among the confirmed Delta and Alpha variants cases were performed on all cumulative outcomes reported as of September 12, 2021. The relative risk (RR), the 95% confidence interval, p value was calculated, and the p value (<0.05) was considered significant for the differentiation between the groups ^8^.

## RESULTS

### UK Delta variant outcomes

We performed analyses of the all the confirmed Delta variant cases in UK (n= 593,572) that were reported as of reporting period ending September 12, 2021. There were 123,620 Delta variant cases reported as of June 21,2021, new cases and outcomes from subsequent reporting periods were obtained by subtracting similar outcomes from prior reporting periods. Table 1 and Table 2 show the distribution of SARS-Cov2 cases, hospitalizations, and deaths among the conformed Delta and Alpha variant cases.

**Table: 1.**
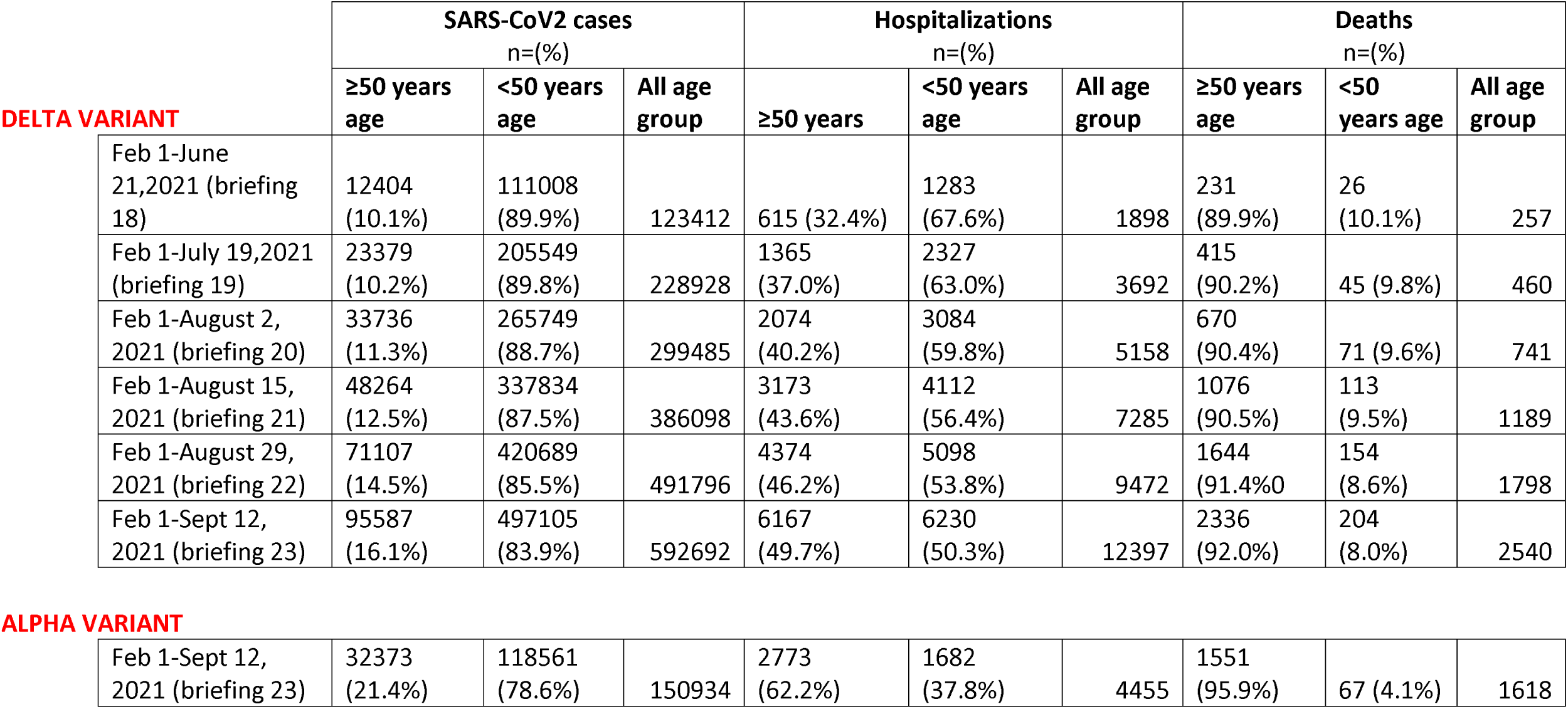
The number of all confirmed (sequencing and genotyping) Delta variant cases, hospitalizations and deaths among age groups as reported from June 21, 2021 to September 12, 2021 reporting periods. The number of all confirmed (sequencing and genotyping) Alpha variant cases, hospitalizations, and deaths as of September 12, 2021 reporting period.

**Table: 2.** Outcomes for all confirmed (sequencing and genotyping) Delta variant SARS-CoV2 cases as reported for each reporting period showing changes in outcomes during the Delta variant surge and outcomes of all confirmed (sequencing and genotyping) Alpha variant cases for the reporting period ending September 12, 2021.

|  | Odds of Hospitalization:<br>hospitalizations/cases (%) |  |  | Hospitalization-death rate:<br>deaths/hospitalizations (%) |  |  | Case Fatality Rate:<br>deaths/cases (%) |  |  |
| --- | --- | --- | --- | --- | --- | --- | --- | --- | --- |
|  | ≥50 years<br>age | <50 years<br>age | All age<br>groups | ≥50 years<br>age | <50<br>years<br>age | All age<br>groups | ≥50 years<br>age | <50 years<br>age | All age<br>groups |
| **DELTA VARIANT** |  |  |  |  |  |  |  |  |  |
| Feb 1-June 21,2021<br>(briefing 18) | 615/1240<br>4 (5.0%) | 1283/1110<br>08 (1.2%) | 1898/12341<br>2 (1.5%) | 231/615<br>(37.6%) | 26/1283<br>(2.0%) | 257/1898<br>(13.5%) | 231/1240<br>4 (1.86%) | 26/11100<br>8 (0.02%) | 257/12341<br>2 (0.21%) |
| Feb 1-July 19,2021<br>(briefing 19) | 1365/233<br>79 (5.8%) | 2327/2055<br>49 (1.1%) | 3692/22892<br>8 (1.6%) | 415/1365<br>(30.4%) | 45/2327<br>(1.9%) | 460/3692<br>(12.5%) | 415/2337<br>9 (1.78%) | 45/20554<br>9 (0.02%) | 460/22892<br>8 (0.20%) |
| Feb 1-August 2,<br>2021 (briefing 20) | 2074/337<br>36 (6.1%) | 3084/2657<br>49 (1.2%) | 5158/29948<br>5 (1.7%) | 670/2074<br>(32.3%) | 71/3084<br>(2.3%) | 741/5158<br>(14.4%) | 670/3373<br>6 (1.99%) | 71/26574<br>9 (0.03%) | 741/29948<br>5 (0.25%) |
| Feb 1-August 15,<br>2021 (briefing 21) | 3173/482<br>64 (6.6%) | 4112/3378<br>34 (1.2%) | 7285/38609<br>8 (1.9%) | 1076/317<br>3 (33.9%) | 113/411<br>2 (2.7%) | 1189/728<br>5 (16.3%) | 1076/482<br>64 (2.23%) | 113/3378<br>34 (0.03%) | 1189/3860<br>98 (0.31%) |
| Feb 1-August 29,<br>2021 (briefing 22) | 4374/711<br>07 (6.2%) | 5098/4206<br>89 (1.2%) | 9472/49179<br>6 (1.9%) | 1644/437<br>4 (37.6%) | 154/509<br>8 (3.0%) | 1798/947<br>2 (19.0%) | 1644/711<br>07 (2.31%) | 154/4206<br>89 (0.04%) | 1798/4917<br>96 (0.37%) |
| Feb 1-Sept 12, 2021<br>(briefing 23) | 6167/955<br>87 (6.5%) | 6230/4971<br>05 (1.3%) | 12397/5926<br>92 (2.1%) | 2336/616<br>7 (37.9%) | 204/623<br>0 (3.3%) | 2540/123<br>97 (20.5%) | 2336/955<br>87 (2.44%) | 204/4971<br>05 (0.04%) | 2540/5926<br>92 (0.43%) |
| **ALPHA VARIANT** |  |  |  |  |  |  |  |  |  |
| Feb 1-Sept 12, 2021<br>(briefing 23) | 2773/323<br>73 (8.6%) | 1682/1185<br>61 (1.4%) | 4455/15093<br>4 (3.0%) | 1551/277<br>3 (55.9%) | 67/1682<br>(4.0%) | 1618/445<br>5 (36.3%) | 1551/323<br>73 (4.79%) | 67/11856<br>1 (0.06%) | 1618/1505<br>61 (1.07%) |

Table 2 shows the percentage of Delta variant cases hospitalized (odds of hospitalizations), percentage of deaths relative to hospitalizations (hospitalization-death rate), and case fatality rate among all age groups, <50 years and ≥50 years old age groups. The overall odds of hospitalization were 1.5%, case fatality rate was 0.21%, hospitalization-death rate was 13.5% of reporting period ending June 21, 2021 for the reporting period ending June 21, 2021 (123,412 cases; 1898 hospitalizations and 257 deaths) among all confirmed Delta variant cases in all age groups. The odds of hospitalization increased to 2.1%, the case fatality rate increased to 0.43% and hospitalization-death rate increased to 20.5% of all the confirmed Delta variant (592,692 cases; 12,397 hospitalizations and 2540 deaths) for the reporting period ending September 12, 2021 in all age groups.

Sub-group analysis ≥50 years age group: The overall case fatality rate for this age group was 1.86%, hospitalization-death rate was 37.6%, and the odds of hospitalization were 5.0% for the reporting period ending June 21, 2021 (12,404 cases, 615 hospitalizations and 231 deaths). The case fatality rate increased to 2.44%, the hospitalization-death rate was 37.9%, and the odds of hospitalization were 6.5% of all Delta variant (95,587 cases; 6167 hospitalizations and 2336 deaths) for the reporting period ending September 12, 2021.

Sub-group analysis <50 years age group: The overall case fatality rate was 0.02%, hospitalizations-death rate was 2.0%, and the odds of hospitalization were 1.2% of the Delta variant (111,008 cases; 1283 hospitalizations and 26 deaths) for the reporting period ending June 21, 2021. The case fatality rate increased to 0.04%, the hospitalization-death rate increased to 3.3%, the odds of hospitalization was 1.3% of all Delta variant (497,105 cases; 6230 hospitalizations and 204 deaths) for the reporting period ending September 12, 2021.

### Outcomes of the confirmed Delta and Alpha variant cases

Table 3 shows the confirmed total Delta variant (592,692 cases; 12,397 hospitalizations and 2540 deaths) and the Alpha variant (150,934 cases; 4455 hospitalizations and 1618 deaths) that were included in the analysis for the reporting period ending September 12, 2021. The Delta variant has a significantly lower overall case fatality rate among all age groups (0.43%vs 1.07%; RR 0.39, P<0.0001); both ≥50 years (2.44% vs 4.79%; RR 0.51, p<0.0001) and <50years age group (0.04% vs 0.06%; RR 0.72, p=0.023) than Alpha variant. The odds of hospitalization are lower among all ages (2.1% vs 3.0%; RR 0.70; P<0.0001), ≥50 years age (6.5% vs 8.6%; RR 0.75; P<0.0001) and <50 years age (1.3% vs 1.4%; RR 0.88; P<0.0001) groups with the Delta variant than the Alpha variant. The hospitalization-deaths rate is significantly lower with Delta variant among ≥50 years (37.9% vs 55.9%; RR 0.67; p<0.0001) than Alpha variant with no significant difference in hospitalization-death rate among <50 years age group (3.3% vs 4.0%, RR 0.82, 95% CI 0.62-1.07, p=0.155). The case fatality rate (0.04% vs 2.44%; RR 0.01; P<0.0001); odds of hospitalization (1.3% vs 6.5%; RR 0.20; P<0.0001) and hospitalization-death rate (3.3% vs 37.9%; RR 0.11; P<0.0001) of <50 years age group is significantly lower than ≥50 years age group among the confirmed Delta variant cases. The case fatality rate; odds of hospitalization and hospitalization-death rate of <50 years age group is significantly lower than ≥50 years age group among the confirmed Alpha variant cases (Table 3).

**Table 3:**
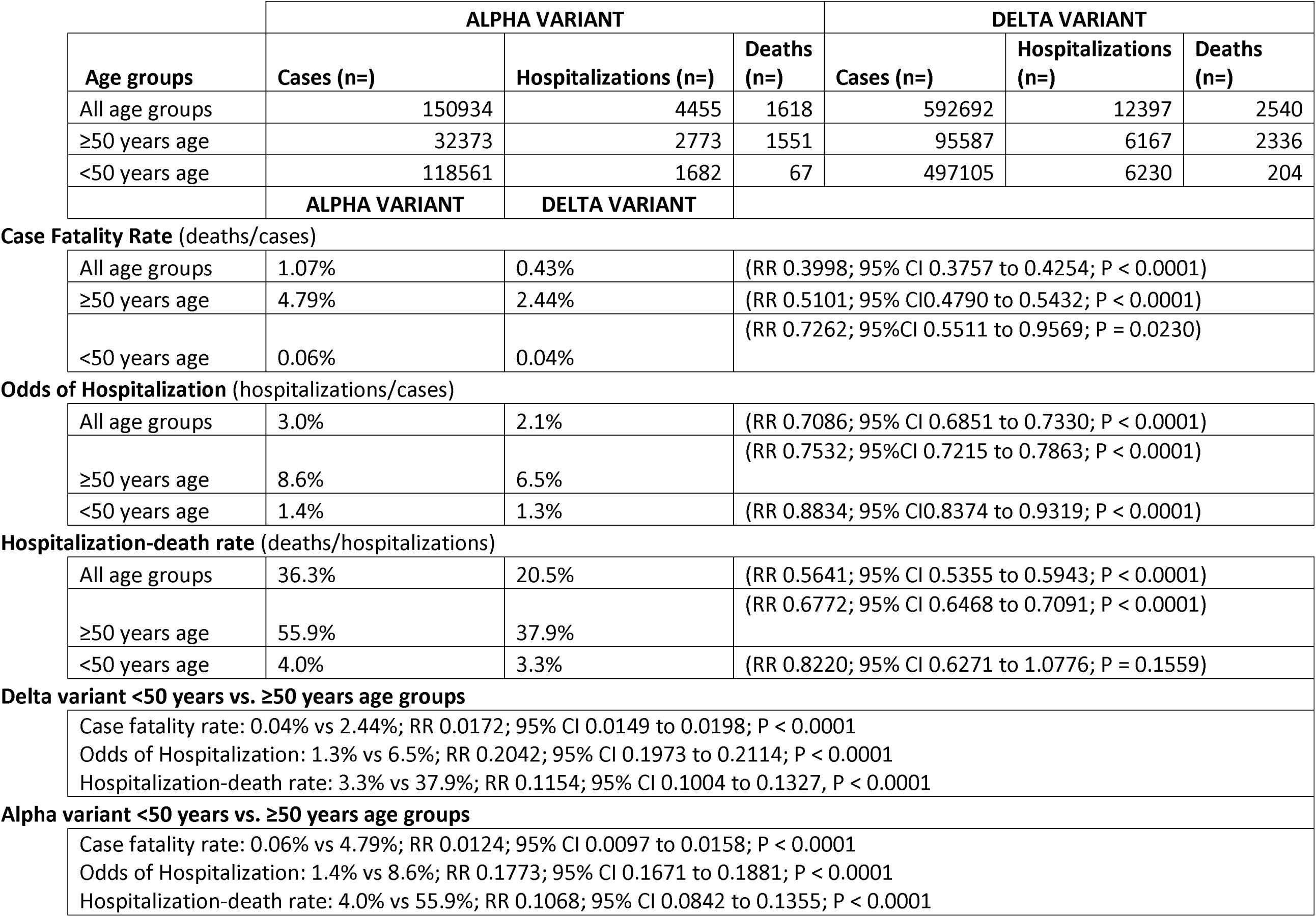
Comparing outcomes of all confirmed (sequencing and genotyping) Delta variant and Alpha variant SARS-CoV2 cases; comparing outcomes <50 years and ≥50 years age groups within Delta or Alpha variant cases as of reporting period ending September 12, 2021.

|  | ALPHA VARIANT |  |  | DELTA VARIANT |  |  |
| --- | --- | --- | --- | --- | --- | --- |
| Age groups | Cases (n=) | Hospitalizations (n=) | Deaths (n=) | Cases (n=) | Hospitalizations (n=) | Deaths (n=) |
| All age groups | 150934 | 4455 | 1618 | 592692 | 12397 | 2540 |
| ≥50 years age | 32373 | 2773 | 1551 | 95587 | 6167 | 2336 |
| <50 years age | 118561 | 1682 | 67 | 497105 | 6230 | 204 |
|  | ALPHA VARIANT | DELTA VARIANT |  |  |  |  |

Case Fatality Rate (deaths/cases)
|  |  |  |  |
| --- | --- | --- | --- |
| All age groups | 1.07% | 0.43% | (RR 0.3998; 95% CI 0.3757 to 0.4254; P < 0.0001) |
| ≥50 years age | 4.79% | 2.44% | (RR 0.5101; 95% CI 0.4790 to 0.5432; P < 0.0001) |
| <50 years age | 0.06% | 0.04% | (RR 0.7262; 95% CI 0.5511 to 0.9569; P = 0.0230) |

Odds of Hospitalization (hospitalizations/cases)
|  |  |  |  |
| --- | --- | --- | --- |
| All age groups | 3.0% | 2.1% | (RR 0.7086; 95% CI 0.6851 to 0.7330; P < 0.0001) |
| ≥50 years age | 8.6% | 6.5% | (RR 0.7532; 95% CI 0.7215 to 0.7863; P < 0.0001) |
| <50 years age | 1.4% | 1.3% | (RR 0.8834; 95% CI 0.8374 to 0.9319; P < 0.0001) |

Hospitalization-death rate (deaths/hospitalizations)
|  |  |  |  |
| --- | --- | --- | --- |
| All age groups | 36.3% | 20.5% | (RR 0.5641; 95% CI 0.5355 to 0.5943; P < 0.0001) |
| ≥50 years age | 55.9% | 37.9% | (RR 0.6772; 95% CI 0.6468 to 0.7091; P < 0.0001) |
| <50 years age | 4.0% | 3.3% | (RR 0.8220; 95% CI 0.6271 to 1.0776; P = 0.1559) |

### Demographic shifts in outcomes during the delta variant surge

As shown on the Table 4, initially during the Delta variant surge as of reporting period ending August 2, 2021, there were 11.3% (n=33,736) of total Delta variant cases (n=299,485), 40.2% (n=2074) of total hospitalizations (n=7239), 90.4% (n=670) of total deaths (n=741) among ≥50years age group. As the Delta variant pandemic progressed there was a significant increased percentage of total cases from 11.3% to 21.1% (RR 1.87; P<0.0001), significantly increased percentage of total hospitalizations from 40.2% to 56.5% (RR 1.40; P<0.001) and trend towards increased percentage total deaths from 90.4% to 92.6% (RR 1.02; P=0.08) among ≥50 years age group confirmed cases of delta variant during August 3-September 12, 2021 period. Table 4 also demonstrate a significantly increased percentage of total COVID-19 deaths occurring among ≥70 years age group [1200 of 1857 (71.4%) vs. 4865 of 6876 (75.1%) total COVID-19 deaths; RR 1.05, P=0028] with 50-69 years old age group showing significant decreased deaths [480 of 1857 (28.6%) vs 1609 of 6876 (24.9%) total COVID-19 deaths; RR 0.86; P=0.0016] during the June 4-July 30, 2021 vs. August 6-October 8, 2021 comparison period.

**Table 4:** Changes in outcomes among demographic ages during the delta variant surge. The confirmed (sequencing and genotyping) SARS-CoV2 Delta variant cases, hospitalizations, and deaths among age groups for period ending June 21, 2021. The outcomes among new reported cases with each subsequent reporting period since immediate prior reporting period until September 12, 2021. There was a significant increase in percent of cases and hospitalizations among confirmed Delta variant cases in ≥50 years old during August 3-September 12, 2021 (week 31-36) period comparison with total cases as of August 2, 2021. Analysis of Total weekly COVID-19 deaths showed significant increase in deaths among ≥50 years old during August 6-October 8, 2021 period on comparison with June 4-July 30, 2021 with most of this significant increase in deaths occurred in over 70 years old age group with 50-69 years age group showing a significant decreased deaths during the same period.

| Confirmed Delta Variant cases from February 1-September 12, 2021 |  |  |  |  |  |  |
| --- | --- | --- | --- | --- | --- | --- |
|  | Cases (%) |  | Hospitalizations (%) |  | Deaths (%) |  |
| *Time period* | ≥50 years age | <50 years age | ≥50 years age | <50 years age | ≥50 years age | <50 years age |
| Feb 1-June 21, 2021 | 12404 (10.1%) | 111008 (89.9%) | 615 (32.4%) | 1283 (67.6%) | 231 (89.9%) | 26 (10.1%) |
| June 22-July 19, 2021 | 10975 (10.4%) | 94541 (89.6%) | 750 (41.8%) | 1044 (58.2%) | 184 (90.6%) | 19 (9.4%) |
| July 20-Aug 2, 2021 | 10357 (14.7%) | 60200 (85.3%) | 709 (48.4%) | 757 (51.6%) | 255 (90.7%) | 26 (9.3%) |
| Aug 3-Aug 15, 2021 | 14528 (16.8%) | 72085 (83.2%) | 1099 (51.7%) | 1028 (48.3%) | 406 (90.6%) | 42 (9.4%) |
| Aug 16-Aug 29, 2021 | 22843 (21.6%) | 82855 (78.4%) | 1201 (54.9%) | 986 (45.1%) | 568 (93.3%) | 41 (6.7%) |
| Aug 30-Sept 12, 2021 | 24480 (24.3%) | 76416 (75.7%) | 1793 (61.3%) | 1132 (38.7%) | 692 (93.3%) | 50 (6.7%) |

| Outcomes of Confirmed Delta variant cases ≥50 years age group |  |  |  |  |  |
| --- | --- | --- | --- | --- | --- |
|  | Feb 1-Aug 2, 2021 |  | Aug 3-Sept 12, 2021 |  |  |
|  | All ages | ≥50 years age | All ages | ≥50 years age |  |
| Cases | 299485 | 33736 (11.3%) | 293207 | 61851 (21.1%) | RR 1.8726; 95% CI 1.8498 to 1.8957; P < 0.0001 |
| Hospitalizations | 5158 | 2074 (40.2%) | 7239 | 4093 (56.5%) | RR 1.4062; 95% CI 1.3525 to 1.4620; P < 0.0001 |
| Deaths | 741 | 670 (90.4%) | 1799 | 1666 (92.6%) | RR 1.0242; 95% CI 0.9971 to 1.0521; P = 0.0806 |

| Total weekly COVID-19 deaths during the Delta Variant surge |  |  |  |
| --- | --- | --- | --- |
|  | June 4-July 30, 2021<br>(week 22-30) | Aug 6-Oct 8, 2021 (Week 31-40) |  |
| **Age group** | 1857 (100%) | 6876 (100%) |  |
| <50 years | 177 (9.5%) | 402 (5.8%) | RR 0.6134; 95 CI 0.5179 to 0.7265; P < 0.0001 |
| ≥50 years | 1680 (90.5%) | 6474 (94.2%) | RR 1.0407; 95% CI 1.0243 to 1.0574; P < 0.0001 |
| 50-69 years | 480 (28.6%) | 1609 (24.9%) | RR 0.8699; 95 CI 0.7977 to 0.9486; P = 0.0016 |
| ≥70 years | 1200 (71.4%) | 4865 (75.1%) | RR 1.0521; 95% CI 1.0176 to 1.0877; P= 0.0028 |

Figures 1-4 show the weekly case rates and weekly hospitalizations per 100,000 population, percentage of weekly deaths among the age groups from July 5, 2020 (week 27) through October 8, 2021 (week 40). The Figure 1 and 3 shows decreased rates of hospitalizations relative to the case rates during the Delta variant surge than the prior Alpha variant surge. Figure 1 demonstrates the increased hospitalizations in younger population during the Delta variant surge peaked during week 29 was associated with 2 to 3 times more weekly percentage of positive cases among younger age groups. This peaked around week 26 and the decreased hospitalization rates since then until the reporting period ending October 8, 2021. Figure 1 also show decreased case rates during the period of summer school break and increased case rates prior to and after summer school break in age groups 5-9 years and 10-19 years. Figure 3 shows gradually increasing proportion of case rates with increasing rates of hospitalizations among ≥55 years age groups.

Figures 2 and 4 show the percentage of total weekly SARS-CoV2 deaths reported since the beginning of the pandemic in early 2020. During the period before the Delta variant surge until May 2021, a substantial percentage of deaths (up to 97.3%) were found to be occurring among ≥50 years age group. In the beginning of June 2021, the percentage of total weekly COVID-19 deaths among ≥50 years went down to 87% with associated increased percentage of total COVID-19 deaths (3.8% weekly deaths in week 34 of 2020 to 10.4% by week 24 in 2021) during the Delta variant surge among 40-49% years old. As the Delta variant surge progressed, by October 13, 2021, the percentage of weekly COVID-19 deaths among ≥50 years increased to 96.7% with most of this increase due to increased deaths among over 70 years old (57.7% at week 21 to 79.5% at week 40 in 2021), with decreased trends in weekly deaths among 60-69 year, 50-59 year and 20-49 year age groups. An increased percentage of total COVID-19 deaths among the over 70 years age group associated with increased proportion of cases during the later part of the Delta variant surge from week 28 until study ending period of week 40 in 2021 (Figures 3-4).

The UK started with phased easing of restrictions (March 2021-phase I, April 2021-Phase 2 reopening) and by May 2021 outdoor social contact were to be removed, two households or six people were allowed to meet indoors, and indoor hospitality services were provided and hotels were opened^22–25^. UK schools were reopened effective March 8, 2021^22^. The UK government announced that most restrictions were lifted in England by July 17, 2021 including face masks mandates, social distancing measures and capacity limitations in venues and these were replaced by recommendations^24,25^.

In summary, of the 593,572 confirmed Delta variant cases that were reported as of September 12, 2021, the Delta variant has an overall lower case fatality rate and lower odds of hospitalization than the Alpha variant. The overall hospitalization-death rate (deaths/hospitalizations) among ≥50 years age group is lower with the Delta variant than the Alpha variant with no statistically significant difference among <50 years age group for the same. The <50 years age group had higher percentage of cases (89.9%) and hospitalization (67.5%) during the beginning of the Delta variant surge with 10.1% of deaths. Despite the lower percentage of cases and hospitalizations during the beginning of pandemic for the reporting period ending August 2, 2021, the ≥50 years age group reported a substantial 90.4% of total COVID-19 deaths. As the Delta variant surge progressed there was a significant increased percentage of cases and hospitalizations occurred among ≥50 years age group confirmed delta variant cases until September 12, 2021. An analysis of the total COVID-19 deaths during the Delta variant surge (June 4-October 8, 2021) showed a significant increased percentage of total COVID-19 deaths occurred among ≥70 years age group during August 6-October 8, 2021 period.

## DISCUSSION

Our analysis of data on the Delta variant in UK showed favorable case fatality rate as was reported in the regular Public Health England briefings during the Delta variant surge^8,15–19^. Our observational study data has shown that the Delta variant is associated with statistically decreased in odds of hospitalization (all ages, <50 years and ≥50 years age groups), and statistically decreased hospitalization death rate among ≥50 years age group than the Alpha variant. These favorable outcomes with the Delta variant relative to the Alpha variant are probably due to less virulence of the Delta variant and association of COVID-19 vaccination among risk groups^26,27^. Our findings defer from an earlier study in the UK that showed higher hospital admissions or emergency care attendance risk for the Delta variant than the Alpha variant infections^9^. However, that study only compared 8,682 Delta variant infections in the beginning of the Delta variant surge and our study involved 593,572 confirmed Delta variant infections outcomes until September 12, 2021 and the outcomes are compared well with total SARS-CoV2 weekly cases, hospitalizations and death in UK until October 8, 2021^9^.

Our data showed comprehensive dynamic changes with decreased percentage of cases, hospitalizations among <50 years and increased percentage of cases and hospitalization among ≥50 years age groups with associated increased case fatality rate and hospitalization death rates during the Delta variant surge (Table 2 and 4, Figures 1-4). Although the younger <50 years age group had the highest proportion of cases and hospitalizations, this age group reported 10.1% of deaths with majority of deaths (89.9%) occurring among ≥50 years age which reported only 10.1% of total cases and 32.4% of total hospitalizations as of June 21, 2021.

The ≥50 years age group had 61.3% of total hospitalizations during the reporting period August 29-September 12, 2021, approaching the Alpha variant high percentage of hospitalizations (62.2%) among ≥50 years age group. There was a significant increased percentage of total COVID-19 deaths noted in ≥70 years old during August 6-October 8, 2021 period compared to June 4-July 30, 2021. This explains in part for the overall increased case fatality rate among the Delta variant cases (0.21% to 0.43%) and overall increased hospitalization-death rate (13.5% to 20.5%) during the course the Delta variant surge in our study.

Our data also clearly showed that the case rates among 5-9 years, 10-19 years age groups are 2-3 folds higher (except some decrease during summer break), and a slight increase in case rates among the 20-29 years old age group during the Delta variant surge than before while all the other age groups showed decreased case rates during the Delta variant surge. This deferential high case rates among younger age groups are most likely due to increased social mobility with behavioral component after step 1-3 of UK reopening with easing of restrictions contributed to increased transmissibility^22–24^. The causes for this increased case rates in younger population due to higher susceptibility of this age groups with Delta variant is a possibility, however the excess case rates can also be explained by increased risk of transmission due to reopening of schools and easing of social restrictions.

COVID-19 vaccination has been shown to be highly effective in reducing symptomatic infections, severe illness and deaths; the improved outcomes during the Delta variant outbreak is probably due to greater adaptation of vaccination among high-risk elderly population^26,27^. However, the Public Health England briefings also show a growing problem of COVID-19 breakthrough cases in the ≥50 years age group, with fully vaccinated share of breakthrough cases increased from 37.0% in briefing 17 to 75.3% of breakthrough cases among fully vaccinated as of September 12, 2021 reporting period in briefing 23. This was associated with 67.0% of subgroup deaths in the fully vaccinated in ≥50 years age group. This was probably due to waning of vaccine efficacy and fully vaccinated high-risk population not taking preventive measures to avoid exposure which need further investigation^8,14–19,28^.

Limitations of our study is, it is an observational study of publicly reported data. However, this database of outcomes among the confirmed 593,572 Delta variant cases in UK technical briefings (as shown in Table 4, for period ending September 12, 2021, week 36), is correlating well with our comprehensive analysis of all the SARS-CoV2 weekly case rates, hospitalizations and deaths during the Delta variant surge in UK for the same period ending week 36 in 2021 (Figures 1-4). The other limitations of our study are the generalizability of the findings is limited to UK population.

## CONCLUSIONS

The <50 years old population had favorable outcomes (significantly lower case fatality rate, odds of hospitalization, and hospitalization-death rates than ≥50 years age group) despite having the highest percentage of cases that contributed to increased hospitalizations initially. Ominous signs of significantly increased percentage of cases and hospitalizations (approaching the Alpha variant proportion of hospitalizations) among ≥50 years age group with associated significantly increased percentage of deaths in this age group especially occurred in ≥70 years of age group.

## Data Availability

All data produced in the present work are contained in the manuscript

## REFERENCES

1. Public Health England. SARS-CoV-2 variants of concern and variants under investigation in England: technical briefing 8. Published online April 1, 2021. Accessed March 31, 2021. https://assets.publishing.service.gov.uk/government/uploads/system/uploads/attachment_data/file/975742/Variants_of_Concern_VOC_Technical_Briefing_8_England.pdf

2. Public Health England. SARS-CoV-2 variants of concern and variants under investigation in England: technical briefing 9. Published online April 22, 2021. Accessed April 21, 2021. https://assets.publishing.service.gov.uk/government/uploads/system/uploads/attachment_data/file/979818/Variants_of_Concern_VOC_Technical_Briefing_9_England.pdf

3. European Centre for Disease Prevention and Control. Threat assessment brief: emergence of SARS-CoV-2 B.1.617 variants in India and situation in the EU/EEA. May 11, 2021. Published online May 11, 2021. Accessed May 10, 2021. https://www.ecdc.europa.eu/sites/default/files/documents/Emergence-of-SARS-CoV-2-B.1.617-variants-in-India-and-situation-in-the-EUEEA_0.pdf

4. Public Health England. SARS-CoV-2 variants of concern and variants under investigation in England: technical briefing 10. Published online May 7, 2021. Accessed May 6, 2021. https://assets.publishing.service.gov.uk/government/uploads/system/uploads/attachment_data/file/984274/Variants_of_Concern_VOC_Technical_Briefing_10_England.pdf

5. Public Health England. SARS-CoV-2 variants of concern and variants under investigation in England: technical briefing 12. Published online May 22, 2021. Accessed June 21, 2021. https://assets.publishing.service.gov.uk/government/uploads/system/uploads/attachment_data/file/988619/Variants_of_Concern_VOC_Technical_Briefing_12_England.pdf

6. Public Health England. SARS-CoV-2 variants of concern and variants under investigation in England: technical briefing 15. Published online June 11, 2021. Accessed June 10, 2021. https://assets.publishing.service.gov.uk/government/uploads/system/uploads/attachment_data/file/993879/Variants_of_Concern_VOC_Technical_Briefing_15.pdf

7. Public Health England. SARS-CoV-2 variants of concern and variants under investigation in England: technical briefing 16. Published online June 18, 2021. Accessed June 17, 2021. https://assets.publishing.service.gov.uk/government/uploads/system/uploads/attachment_data/file/1001359/Variants_of_Concern_VOC_Technical_Briefing_16.pdf

8. Public Health England. SARS-CoV-2 variants of concern and variants under investigation in England: technical briefing 23. Published online September 17, 2021. Accessed September 16, 2021. https://assets.publishing.service.gov.uk/government/uploads/system/uploads/attachment_data/file/1018547/Technical_Briefing_23_21_09_16.pdf

9. Twohig KA, Nyberg T, Zaidi A, et al. Hospital admission and emergency care attendance risk for SARS-CoV-2 delta (B.1.617.2) compared with alpha (B.1.1.7) variants of concern: a cohort study. The Lancet Infectious Diseases. Published online August 2021. doi:10.1016/S1473-3099(21)00475-8

10. New York Times. Hospitalizations for children sharply increase as Delta surges, C.D.C. studies find. Published online 2021. Accessed September 2, 2021. https://www.nytimes.com/2021/09/03/health/delta-children-hospitalization-rates.html

11. Healthline. Young People Make Up Biggest Group of Newly Hospitalized COVID-19 Patients. Published online 2021. Accessed July 28, 2021. https://www.healthline.com/health-news/young-people-make-up-biggest-group-of-newly-hospitalized-covid-19-patients

12. CDC: Morbidity and Mortality Weekly Report (MMWR). Severity of Disease Among Adults Hospitalized with Laboratory-Confirmed COVID-19 Before and During the Period of SARS-CoV-2 B.1.617.2 (Delta) Predominance — COVID-NET, 14 States, January–August 2021. Published online 2021. Accessed October 21, 2021. https://www.cdc.gov/mmwr/volumes/70/wr/mm7043e1.htm?s_cid=mm7043e1_w

13. CDC: Morbidity and Mortality Weekly Report (MMWR). Distribution of SARS-CoV-2 Variants in a Large Integrated Health Care System — California, March–July 2021. Published online 2021. Accessed October 7, 2021. https://www.cdc.gov/mmwr/volumes/70/wr/mm7040a4.htm

14. Public Health England. SARS-CoV-2 variants of concern and variants under investigation in England: technical briefing 17. Published online June 25, 2021. Accessed June 24, 2021. https://assets.publishing.service.gov.uk/government/uploads/system/uploads/attachment_data/file/1001354/Variants_of_Concern_VOC_Technical_Briefing_17.pdf

15. Public Health England. SARS-CoV-2 variants of concern and variants under investigation in England: technical briefing 18. Published online July 9, 2021. Accessed July 8, 2021. https://assets.publishing.service.gov.uk/government/uploads/system/uploads/attachment_data/file/1001358/Variants_of_Concern_VOC_Technical_Briefing_18.pdf

16. Public Health England. SARS-CoV-2 variants of concern and variants under investigation in England: technical briefing 19. Published online July 23, 2021. Accessed July 22, 2021. https://assets.publishing.service.gov.uk/government/uploads/system/uploads/attachment_data/file/1005517/Technical_Briefing_19.pdf

17. Public Health England. SARS-CoV-2 variants of concern and variants under investigation in England: technical briefing 20. Published online August 6, 2021. Accessed August 5, 2021. https://assets.publishing.service.gov.uk/government/uploads/system/uploads/attachment_data/file/1009243/Technical_Briefing_20.pdf

18. Public Health England. SARS-CoV-2 variants of concern and variants under investigation in England: technical briefing 21. Published online August 20, 2021. Accessed August 19, 2021. https://assets.publishing.service.gov.uk/government/uploads/system/uploads/attachment_data/file/1012644/Technical_Briefing_21.pdf

19. Public Health England. SARS-CoV-2 variants of concern and variants under investigation in England: technical briefing 22. Published online September 3, 2021. Accessed September 2, 2021. https://assets.publishing.service.gov.uk/government/uploads/system/uploads/attachment_data/file/1014926/Technical_Briefing_22_21_09_02.pdf

20. Public Health England. National flu and COVID-19 surveillance reports. Published online 2021. Accessed October 15, 2021. https://www.gov.uk/government/statistics/national-flu-and-covid-19-surveillance-reports

21. UK office of National Statistics. Deaths registered weekly in England and Wales, provisional. Published online 2021. Accessed October 6, 2021. https://www.ons.gov.uk/peoplepopulationandcommunity/birthsdeathsandmarriages/deaths/datasets/weeklyprovisionalfiguresondeathsregisteredinenglandandwales/2021

22. UK Prime Minister’s Office. Schools and colleges to reopen from tomorrow as part of Step One of the roadmap. Published online 2021. Accessed March 6, 2021. https://www.gov.uk/government/news/schools-and-colleges-to-reopen-from-tomorrow-as-part-of-step-one-of-the-roadmap

23. UK Prime Minister’s Office. Further easing of Covid restrictions confirmed for 12 April -Outdoor pubs, shops, gyms and hairdressers to reopen. Published online 2021. Accessed April 4, 2021. https://www.gov.uk/government/news/further-easing-of-covid-restrictions-confirmed-for-12-april

24. UK Prime Minister’s Office. Further easing of COVID restrictions confirmed for 17 May -Planned Step 3 easements will go ahead on 17 May. Published online 2021. Accessed May 9, 2021. https://www.gov.uk/government/news/further-easing-of-covid-restrictions-confirmed-for-17-may

25. UK Prime Minister’s Office. Prime Minister to set out plans ahead of step 4. Published online 2021. Accessed July 4, 2021. https://www.gov.uk/government/news/prime-minister-to-set-out-plans-ahead-of-step-4

26. Lopez Bernal J, Andrews N, Gower C, et al. Effectiveness of the Pfizer-BioNTech and Oxford-AstraZeneca vaccines on covid-19 related symptoms, hospital admissions, and mortality in older adults in England: test negative case-control study. BMJ. Published online May 13, 2021. doi:10.1136/bmj.n1088

27. Lopez Bernal J, Andrews N, Gower C, et al. Effectiveness of Covid-19 Vaccines against the B.1.617.2 (Delta) Variant. New England Journal of Medicine. Published online July 21, 2021. doi:10.1056/NEJMoa2108891

28. Emani VR, Reddy R, Goswami S. Effectiveness of Covid-19 Vaccines against the B.1.617.2 (Delta) Variant. New England Journal of Medicine. 2021;385(25):e92. doi:10.1056/NEJMc2113090

